# Deep Survival Analysis for Interpretable Time-Varying Prediction of Preeclampsia Risk

**DOI:** 10.1101/2024.01.18.24301456

**Authors:** Braden W Eberhard, Kathryn J Gray, David W Bates, Vesela P Kovacheva

## Abstract

**Objective:** Survival analysis is widely utilized in healthcare to predict the timing of disease onset. Traditional methods of survival analysis are usually based on Cox Proportional Hazards model and assume proportional risk for all subjects. However, this assumption is rarely true for most diseases, as the underlying factors have complex, non-linear, and time-varying relationships. This concern is especially relevant for pregnancy, where the risk for pregnancy-related complications, such as preeclampsia, varies across gestation. Recently, deep learning survival models have shown promise in addressing the limitations of classical models, as the novel models allow for non-proportional risk handling, capturing nonlinear relationships, and navigating complex temporal dynamics.

**Methods:** We present a methodology to model the temporal risk of preeclampsia during pregnancy and investigate the associated clinical risk factors. We utilized a retrospective dataset including 66,425 pregnant individuals who delivered in two tertiary care centers from 2015-2023. We modeled the preeclampsia risk by modifying DeepHit, a deep survival model, which leverages neural network architecture to capture time-varying relationships between covariates in pregnancy. We applied time series k-means clustering to DeepHit’s normalized output and investigated interpretability using Shapley values.

**Results:** We demonstrate that DeepHit can effectively handle high-dimensional data and evolving risk hazards over time with performance similar to the Cox Proportional Hazards model, achieving an area under the curve (AUC) of 0.78 for both models. The deep survival model outperformed traditional methodology by identifying time-varied risk trajectories for preeclampsia, providing insights for early and individualized intervention. K-means clustering resulted in patients delineating into low-risk, early-onset, and late-onset preeclampsia groups— notably, each of those has distinct risk factors.

**Conclusion:** This work demonstrates a novel application of deep survival analysis in time-varying prediction of preeclampsia risk. Our results highlight the advantage of deep survival models compared to Cox Proportional Hazards models in providing personalized risk trajectory and demonstrating the potential of deep survival models to generate interpretable and meaningful clinical applications in medicine.

## 1. Introduction

Precise modeling of disease risk is critical in healthcare and serves diverse purposes, such as enabling improved clinical decision-making, predicting time-to-event outcomes, advancing precision medicine, and refining disease classification strategies[1–3]. Time-to-event or survival analysis can provide meaningful insights for understanding temporal disease dynamics. While traditional parametric and semiparametric models, such as the Cox Proportional Hazards model [4], have been effective in analyzing disease outcomes, their assumption of constant hazard ratios over time may be poorly suited for diseases limited to specific life periods, such as pregnancy-related conditions. Furthermore, the Cox model’s sensitivity to censored data and assumption of covariate linearity can introduce bias and potentially oversimplify the temporal dynamics of disease relationships.

In recent years, deep learning techniques have transformed survival analysis in healthcare, yielding improvements across various categories of models. Time-dependent models, such as Cox-Time[5], exemplify this advancement, with an enhanced capacity to handle dynamic predictors, addressing a limitation of traditional methods by accommodating features that change over time. High-dimensional models, including DeepConvSurv[6] and CapSurv[7], showcase the newfound ability to integrate and interpret information from diverse data modalities, such as images, using convolutional neural networks. Furthermore, the incorporation of deep learning in parametric models, as observed in DeepPAMM[8] and DPWTE[9], advances survival analysis frameworks that leverage probabilistic models for more accurate predictions. DeepHit[10] introduces an innovative discrete-time approach to manage time-to-event data through classification techniques. This model excels in adapting to time-varying effects without making assumptions about the underlying hazard risk distribution within a high-dimensional space. This provides additional flexibility in scenarios where traditional models encounter challenges and enhances the model’s inherent ability to capture nonlinear relationships. These advancements collectively highlight the impact of deep learning, resulting in models with heightened predictive power, augmented interpretability, increased flexibility, and broader applicability in healthcare settings.

Preeclampsia, a hypertensive disorder specific to pregnancy, is a major cause of severe maternal and neonatal morbidity and mortality with an incidence of 2-8%[11][12]. Preeclampsia is defined by new-onset high blood pressure and proteinuria after 20 weeks of gestation and can lead to end-organ damage, including seizures, renal failure, and even death if left untreated[13]. In the United States, preeclampsia rates increased by 25% between 1987 and 2004[14]. The disease may have a distinct biochemical signature and clinical course depending on the specific patient and the time in pregnancy during which the symptoms develop[15]. Early-onset preeclampsia starts before 34 gestational weeks and commonly follows a severe course. As there is no definitive cure other than delivery, preeclampsia that develops early in pregnancy is one of the most common causes of severe maternal morbidity, iatrogenic preterm birth, and associated neonatal complications[16]. Late-onset preeclampsia, characterized by disease onset after 34 gestational weeks or in the postpartum period, tends to have a milder progression and more favorable outcomes. The current clinical practice focuses on identifying at-risk individuals based on clinical criteria, increased surveillance, and early diagnosis and may fail to identify up to 50% of those at risk. In those at high risk, aspirin administration prior to 16 gestational weeks is associated with more favorable outcomes[17]. Earlier identification of at-risk patients, especially those with a high risk of early-onset preeclampsia, would allow for timely prophylaxis and increased surveillance.

Current methods approach preeclampsia risk prediction by reducing the task to a classification problem[18]. Despite their overall accurate performance, most models are limited in predicting disease timing, providing long-term forecasts, or differentiating among antepartum, intrapartum, and postpartum risk[19,20]. More recent methods incorporate competing risk models[21], which allow a probabilistic prediction over time. These models are based on the underlying assumption of Gaussian distribution for risk and do not consider the non-exclusive relationship between birth and preeclampsia, as the disease develops postpartum in up to 27.5% of all pregnancies [22]. Prior studies have used the Cox proportional hazards model in pregnancy to identify hazard ratios in a limited number of features by using gestational age as the outcome and relied on the assumptions of homogenous risk and linear covariate relationships[15].

Our novel approach to addressing preeclampsia onset risk involves a departure from prevailing models, with a fundamental shift of applying survival analysis techniques. While typical survival analysis applications account for right-censored patients, the finite nature of pregnancy ensures the participation of all at-risk patients for the duration of the study. This contrasts with models for other medical conditions, such as heart failure or cancer [1–3], where patient attrition may skew results. Our application in pregnancy-related conditions has several substantial advantages. The temporal influences inherent to conditions, such as preeclampsia, including the critical factors of gestational age and delivery timing, integrate well with the methodology of deep survival models. The varied risk factors and clinical consequences linked to the antepartum, intrapartum, and postpartum manifestations of preeclampsia[23] emphasize the need for a model adept at discerning these time-varying disease onsets. This yields a profile that significantly deviates from conventional parametric distributions.

To address the challenges posed by the temporal relationship of pregnancy and preeclampsia, we implement DeepHit, a specialized deep learning model for survival analysis in healthcare, tailored to predict time-to-event outcomes[10]. Our modification of the competing-risk DeepHit architecture conforms the model to predict a single event –preeclampsia diagnosis. Importantly, it allows the model to handle hazard functions nonparametrically to reflect the nonproportional risk across different stages of pregnancy. This enhanced flexibility allows DeepHit to predict disease risk and discriminate between distinct temporal disease risk trajectories. Through these advancements, our model seeks to provide a more accurate understanding of time-varied preeclampsia risk, enhancing the precision and depth of predictive capabilities in the complex landscape of maternal health.

To comprehensively evaluate the performance of our approach, we conducted a comparative analysis between DeepHit and a traditional semiparametric Cox proportional hazards model[4], which served as a baseline model. This comparative study underscores the distinctive features and advantages offered by our personalized deep learning approach. Although the Cox proportional hazards model has been a reliable tool for accurately portraying disease prognosis, its inherent dependence on proportional hazards limits its ability to adjust to the time-varying disease risk profiles frequently encountered in real-world clinical scenarios, such as preeclampsia[24]. This inherent limitation of the Cox model guarantees that the risk for each patient maintains a consistent proportionality to a baseline risk function throughout the entire observational period.

We present the overall predictive accuracy of DeepHit to facilitate a comparative evaluation with other classification models within the same category. Additionally, we leverage time series clustering to unveil distinct patient trajectories, providing a nuanced understanding of preeclampsia progression. Employing explainability techniques emphasizes the model’s clinical relevance: facilitating clinician trust, integration into decision-making, and offering avenues for future research. The outline of our deep survival analysis pipeline is summarized in Fig. 1. Our approach aims to extend conventional models by offering a comprehensive perspective on time-to-event predictions in disease risk, particularly in capturing complex temporal dynamics, non-linear relationships, and time-varying risk trajectories in the context of preeclampsia.

**Fig 1.**
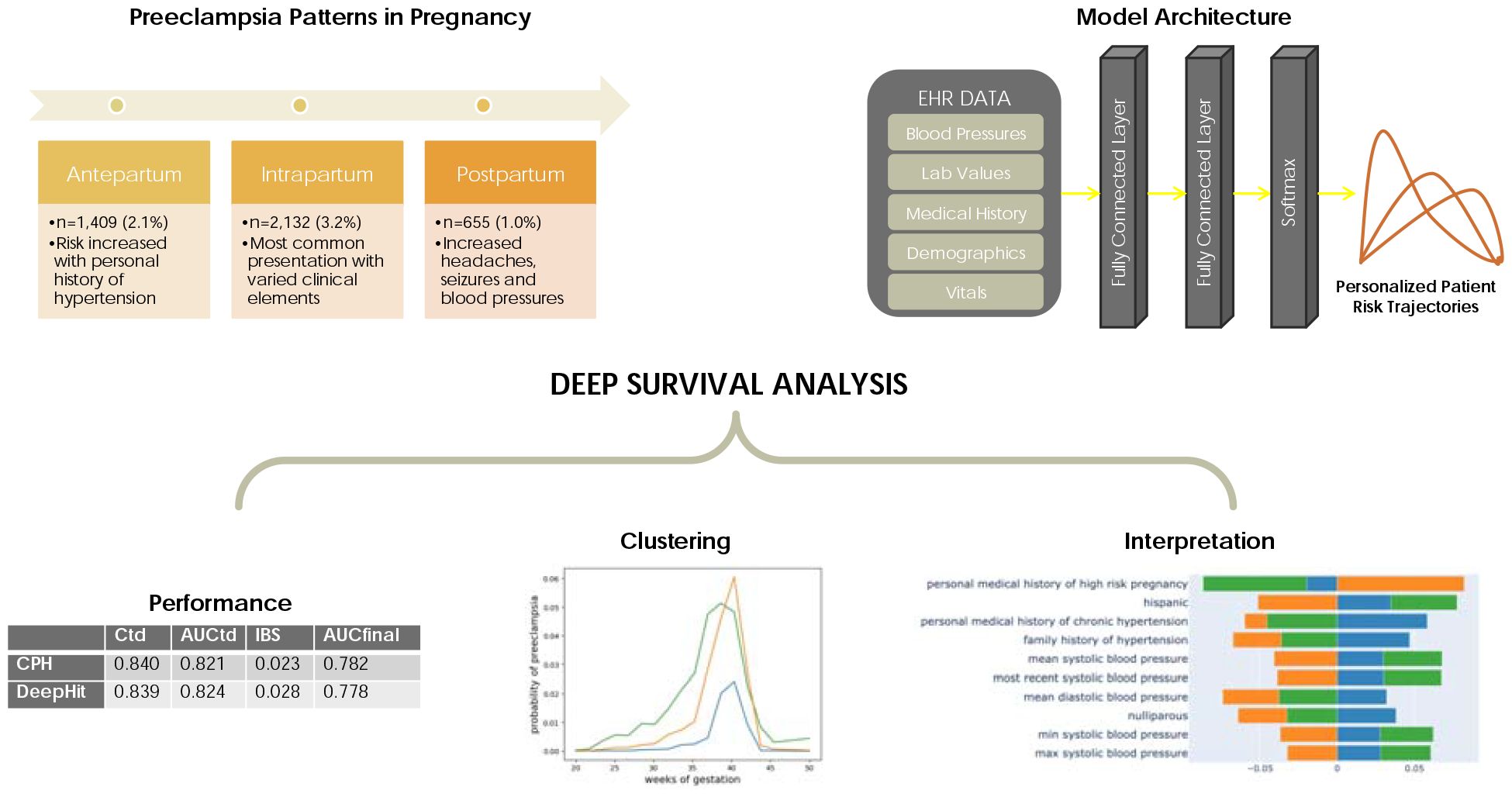
Outline of the deep survival pipeline. This novel approach allows for personalized and interpretable risk prediction of disease onset. Figure 1. Results from the deep survival analysis. A) Time series k-means clustering with DeepHit B) Time series k-means clustering with Cox Proportional Hazards model C) Kaplan Meier survival by DeepHit clusters D) Average preeclampsia risk function by DeepHit clusters. Abbreviations, CPH: Cox Proportional Hazards model

## 2. Material and methods

### 2.1 Dataset

This study was approved by the Mass General Brigham Institutional Review Board, protocol # 2020P002859, with a waiver of patient consent. Selection criteria for pregnant patients were documented pregnancy beyond 20 weeks of gestation and associated billing codes for cesarean or vaginal delivery. We included all available patients from May 2015 to May 2023 and analyzed each pregnancy independently. These dates were chosen as May 2015 is when our institution implemented electronic health records across all outpatient offices and inpatient sites. All data (including sociodemographic, clinical diagnoses, laboratory, and vital signs) were obtained and analyzed using our machine learning platform, which extracts, transforms, and harmonizes data from multiple sources[25]. Several features, including blood pressure, heart rate, weight, and laboratory values, often contained multiple readings. We engineered multiple features to capture how these values change over time, such as minimal, maximal, and range over defined periods of time. As many of these features were highly collinear, only the most clinically relevant were used in the Cox model as assessed using the variance inflation factor[26], such as maximum blood pressure and minimum platelet value. The available features and those included in the models are reported in Supplementary Tables 1 and 2, respectively.

We included only patient data obtained up to 20 gestational weeks to minimize data leakage risk and generate meaningful clinical insights. By definition, the outcome is unavailable before 20 gestational weeks[12], and the disease is rarely diagnosed before 24 gestational weeks. This cutoff was also selected to ensure the clinical relevance of the predictions since aspirin, a prophylactic medication that can decrease disease severity, especially for early-onset preeclampsia, exerts optimal prophylactic effects if started before 20 weeks[17]. We defined the outcome, preeclampsia, as the presence of hypertension and proteinuria after 20 gestational weeks in a previously normotensive individual, based on the established guidelines[12]. The time stamp associated with the diagnosis was the first instance when the diagnostic criteria were met.

The study involved a cohort of 52,298 patients and 66,425 deliveries; each pregnancy was considered an independent event. Within this cohort, there were 4,196 pregnancies with a diagnosis of preeclampsia, representing 6.3% of all deliveries. Within the preeclampsia group, 1,409 cases were diagnosed antepartum, 2,132 intrapartum, and 655 postpartum, accounting for 33.6%, 50.8%, and 15.6% of the cases, respectively (Supplementary Fig. 1). The cohort was diverse, with 63% of the patients had self-reported White race; 19% of individuals identified as Hispanic. The prevalence of preeclampsia diagnoses differed significantly between births before 37 weeks of gestation (32%) and full-term births (7%) (*chi* — *squared* test, p < 0.01). Additional cohort characteristics are detailed in Table 1.

**Table 1.** Cohort characteristics

|  | **Preeclampsia**<br>**(n=4196)** | **No preeclampsia**<br>**(n=62,229)** |
| --- | --- | --- |
| Self-Reported Race |  |  |
| White | 2380 (57 %) | 40002 (64 %) |
| Black | 815 (19 %) | 6652 (11 %) |
| Other | 1029 (25 %) | 15903 (26 %) |
| Hispanic | 855 (20 %) | 10510 (17 %) |
| Non-Hispanic | 3341 (80 %) | 51719 (83 %) |
| Brigham and Women’s Hospital | 2573 (61 %) | 39512 (63 %) |
| Mass General Hospital | 1623 (39 %) | 22717 (37 %) |
| Nulliparous | 820 (20%) | 9378 (15%) |
| Gestational age at delivery, weeks | 37.3 (35.9 - 39.0) | 39.3 (38.4 - 40.1) |
| Last BMI before pregnancy, kg/m<sup>2</sup> | 28.2 (24.0 – 33.8) | 24.8 (22.0 – 29.0) |
| Maximal SBP during pregnancy, mmHg | 175 (163 – 185) | 140 (131 – 149) |
| Maximal DBP during pregnancy, mmHg | 104 (98 – 111) | 86 (81 – 93) |
| Family history of hypertension | 3205 (76 %) | 42654 (69 %) |
| Abbreviations: BMI, body mass index, SBP, systolic blood pressure, DBP, diastolic blood pressure |  |  |

### 2.2. Survival analysis

Typical survival data provide three parts for each observation: 1) covariate features, 2) time elapsed since the start of observation, and 3) a label indicating the occurrence or non-occurrence of the event. We consider survival time discrete intervals with a predetermined maximum time horizon *T*_max_ resulting in a time set of *τ* = {0, …, *T*_max_} where *T*_max_ = 50 weeks. While most survival analysis includes right-censored data due to patients being lost to follow-up, this exclusively occurs outside our window of observation because preeclampsia is a condition that can only be diagnosed during pregnancy and up to six weeks postpartum. We consider *K*=1 events, specifically the diagnosis of preeclampsia within the time horizon. All patients either have a diagnosis recorded or reach the maximum time denoted by Ø, thus the set of events is *κ* = {Ø, 1}. Each patient is represented by a triple (x,*t,k*) were x ∈ *X* is a *D*-dimensional vector of covariate features, *t*∈ *τ* is either time of diagnosis or *T*_max_, and *k* ∈ *κ* is reflects whether the patient received a diagnosis or not within the time of observation. We are subsequently given a dataset 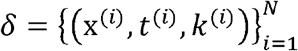 that describes a set of observed patients. Our task is to find 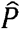, the estimate of the true probability that a patient with covariate features x* will be diagnosed at time *t*\* given by *P*(*t =t*\*,*k* = *k*^*\**^|x = x*).

#### 2.2.1. Cox proportional hazards model for survival analysis

In the Cox proportional hazards model, the hazard function *h*(*t*,X) expresses the probability that a subject who is normotensive at time *t* will be diagnosed at time *t + Δt*. It is defined as the product of a baseline hazard function *h*_O_ (*t*) and an exponential function exp (*β*X) involving covariates X and coefficients β, where *t* denotes time as shown in the formula below.

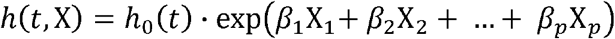

To estimate the regression coefficients *β*_1_,*β*_2_ … *β*_*p*_ in the Cox proportional hazards model, the method of partial likelihood is employed. The partial likelihood is defined as:

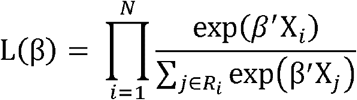

The partial likelihood function, denoted as L(*β*_1_), encapsulates the information regarding the ordering of event times while circumventing the necessity of specifying the baseline hazard function *h*_O_(*t*). Here, denotes the total number of patients with preeclampsia, *R*_*i*_ is the risk set for the *i*-th event or in other words the set of individuals for whom is without previous diagnosis and is still relevant, and *β*’X_*i*_ represents the linear predictor for individual *i*. Model fitting involves the maximization of the partial likelihood, often achieved through iterative optimization algorithms. The log-partial likelihood, denoted as 𝓁(*β*) is utilized for numerical stability and computational efficiency:

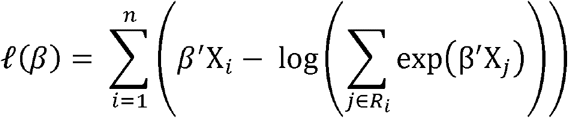

Minimizing the negative log-partial likelihood is equivalent to maximizing the partial likelihood and yields the maximum partial likelihood estimates for the regression coefficients. This process ensures optimal fitting of the Cox proportional hazards model to the observed time-to-event data, enabling robust inference regarding the impact of covariates on the hazard function. The model assumes that hazard ratios remain constant over time, mathematically expressed as:

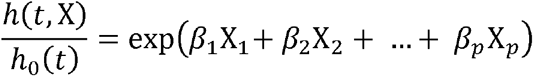

This assumption ensures the proportional impact of covariates on the hazard function.

#### 2.2.2 DeepHit model architecture

The DeepHit model[10] employs an architecture that learns the joint distribution of survival times and competing events by utilizing a shared sub-network and event-specific sub-networks for each event. We modified our network to capture the marginal distribution of preeclampsia risk to address the independent nature of delivery and diagnosis as nonexclusive events. Our implementation takes advantage of shared subnets to pass information freely between all predictions without the competing risks of multiple subnetworks. We minimize a single event loss function of the sum of two terms. The first expression encapsulates the data contributed by diagnosed patients, while the second term leverages the presence of patients who remain undiagnosed at the end of observation. ℒ_*log-likelihood*_ is the negative log-likelihood of the marginal distribution of the first hitting time and event, as shown below.

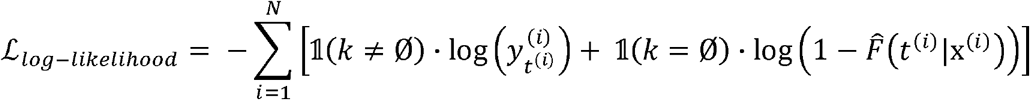

Where *y*_*t*_ is the probability that the patient will develop preeclampsia at time *t*. The indicator function 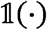 and indicates if the patient was diagnosed (1) or remained healthy(Ø) at the end of observation. We use the estimate cumulative incidence function (CIF) 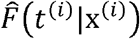 to illustrate the risk of an event occurring where 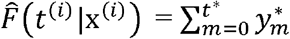

ℒ_*CIF*_ incorporates the estimated cumulative incidence function that diagnosis will occur on or before time *t*, conditional on covariates x calculated at each time. ℒ_*CIF*_ utilizes a ranking loss function which adapts the idea of concordance[27]: a patient who develops the disease at time s should have a higher risk at time *t* than a patient who was disease-free longer than *t*. We adjust ℒ_*CIF*_ for single event risk defined as

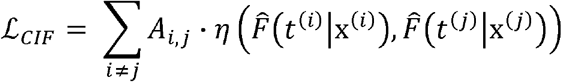

Where *η*(*x,y*) is a convex loss function and 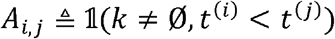. Adjusted for single-event risk, ℒ_*CIP*_ penalizes incorrect ordering of pairs for each patient with a preeclampsia diagnosis. Incorporating this loss into the total loss function expresses the probability that the preeclampsia diagnosis occurs on or before time *t*.

### 2.3 Model training

For evaluation purposes, we split the data into training set (60%), testing set (20%) and validation set (20%). We used Optuna[28], an open-source hyperparameter optimization framework that automates the search for optimal parameters in machine learning models, to search the parameter space and compared discriminative performance on the testing set to select optimal hyperparameters. The final model included 12 layers, each with 128 nodes, combined with ReLu activation functions over 16 distinct time intervals. We trained the network using the Adam optimizer with a batch size of 350 and a learning rate of 0.01. To mitigate overfitting, early stopping was performed based on the loss on the testing data after 8 non-improving iterations. To address class imbalance and enhance the model’s generalizability, oversampling the minority class was implemented resulting in each positive case being represented twice in the training data.

### 2.4 Model Evaluation

In addition to metrics to examine the predictive performance of DeepHit, we employ clustering and explainability methods. These techniques provide insight into the model’s ability to temporally discriminate and uncover underlying patterns within the data, augmenting our understanding of the model’s predictive dynamics.

#### 2.4.1 Predictive Metrics

In this study, the primary metric employed is the time-dependent Concordance Index (Ctd), serving as a key measure to assess the agreement between the ordering of predicted risk and actual survival times. We define Ctd as the proportion of patients that are correctly identified as not having been diagnosed with preeclampsia summed over each time interval:

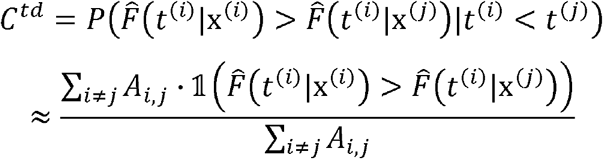

The Integrated Brier Score (IBS) is another performance metric chosen to evaluate the overall accuracy of predicted survival probabilities throughout a specified time interval. By considering the entire survival curve, the Integrated Brier Score quantifies the average squared difference between predicted probabilities and observed outcomes. This metric offers a comprehensive assessment of the model’s performance in estimating survival probabilities over time.

Mathematically, the Integrated Brier Score is the sum of the estimated CIF minus the observed CIF squared:

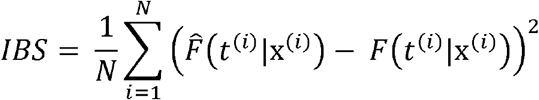

Time-dependent Area Under the Receiver Operating Characteristic Curve (AUCtd)[29] is incorporated to gauge the discriminatory power of the model across each time point in the survival process. This dynamic metric provides a time-varying measure of classification accuracy, capturing changes in predictive performance over the course of the study. AUCtd is the average AUC calculated for each time interval.

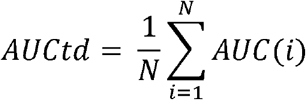

Moreover, we assess the model’s overall prediction of preeclampsia risk across the pregnancy window by computing the Area Under the Receiver Operating Characteristic Curve (AUC) for the final survival probability. This metric, referred to as AUCfinal, allows for a comparison with conventional classification models, providing a comprehensive evaluation of the model’s predictive capacity. Using AUCfinal, we enhance our ability to understand the model’s performance in predicting the overall preeclampsia risk.

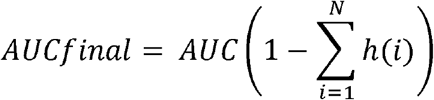

#### 2.4.2 Time Series Clustering

The K-means clustering, a well-established unsupervised learning algorithm, has been adapted and extended to accommodate time series, allowing for the identification of meaningful patterns and trends over time. This method involves representing time series as vectors, with subsequent application of K-means clustering techniques to effectively group similar temporal patterns[30].

Prior to clustering, a normalization procedure is implemented to ensure a comprehensive analysis that captures diverse risk profiles, as opposed to a singular focus on overall risk. Specifically, the hazards of each patient are normalized within the range of 0 to 1 prior to the clustering process. This normalization addresses variations in overall patient risk, allowing for a grouping based on temporal patterns independently of global risk measures.

During clustering, the distance between a patient’s hazard function *x*^(*i*)^ and cluster centroid Cluster *c*^(*j*)^ is quantified through a normalized Euclidean distance metric. This metric, is minimized during the clustering process, thereby optimizing the segregation of temporal patterns within the dataset.

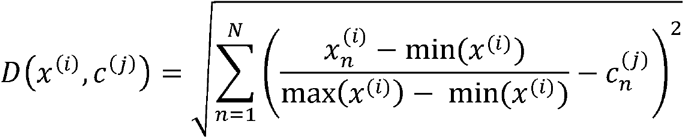

The incorporation of normalization and time series clustering enhances the capacity to discern groups of patients with diverse risk evolutions over time. These trajectories may offer valuable insights into disease presentation, timing, and associated risk. This nuanced approach allows for a more granular understanding of temporal patterns within the data, contributing to personalized interpretation of patient outcomes.

#### 2.4.3 Interpretability

SHAP values, often denoted as SHAP (SHapley Additive exPlanations) values elucidate a model’s output by attributing contributions to each feature in a given prediction. Grounded in cooperative game theory, SHAP values represent the average marginal contribution of each feature across all possible combinations, ensuring an equitable distribution of credit for the model’s output among individual features. In selecting feature importance methods, we opted for SHAP over alternatives like LIME and other methods due to its demonstrated superiority in unsupervised environments and enhanced discriminative power[31]. We calculated SHAP values for the integrated DeepHit and time series clustering model, thus ensuring that the SHAP values represent the differences among clusters generated by DeepHit rather than the cumulative effect on risk prediction. The incorporation of SHAP value analysis is instrumental in showcasing the diverse applications of DeepHit within the context of risk trajectories. By leveraging SHAP values, we demonstrate how each feature influences a particular prediction, contributing to a more comprehensive interpretation of DeepHit’s predictive capabilities and fostering transparency in its risk assessment.

## 3. Results

### 3.1 Model Evaluation Metrics

Evaluation metrics for models on the validation dataset are shown in Table 2. Notably, both Cox proportional hazards and DeepHit models exhibit comparable and effective predictive capabilities, as evidenced by their Ctd, AUCtd, and IBS scores. The close alignment of these metrics underscores the robustness of both models in capturing the temporal dynamics of preeclampsia risk, providing a solid foundation for their reliability in predicting individualized risk trajectories. Moreover, the AUCfinal indicates accurate overall risk prediction and comparable performance between DeepHit and Cox proportional hazards models in a way that allows for meaningful comparisons with classification models.

**Table 2.**
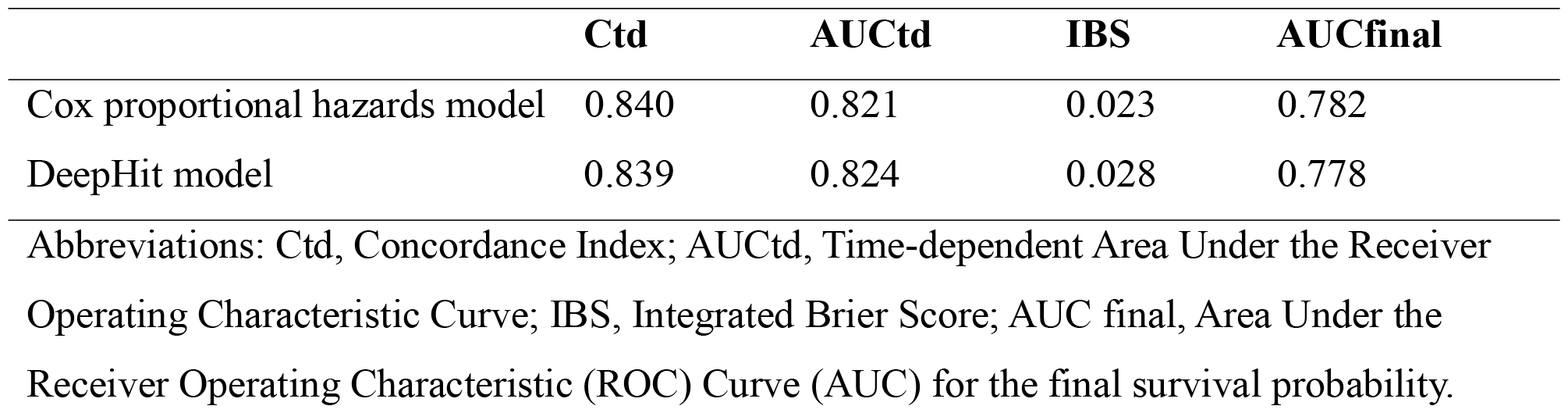
Evaluation metrics on the validation cohort

### 3.2 Clustering by Patient Trajectory

The time series clustering on DeepHit outputs, depicted in Fig. 1 A, highlights the advantages of employing nonparametric models. This method segregates patients into three distinct clusters: low-risk, early preeclampsia, and late preeclampsia. Such clustering adeptly captures variations in risk trajectories, emphasizing the superiority of nonparametric models in discerning nuanced shifts in risk dynamics over time. Notably, the survival outcomes for preeclampsia within these clusters exhibit significant differences, as evident from the log-rank test (p < 0.001 for all comparisons) on Kaplan-Meier estimates shown in Fig. 1C and the mean risk distributions in Fig. 1D. Due to the proportional hazards’ assumption, Cox proportional hazards models’ clustering tends to reflect overall risk rather than individual time-varying risk (Fig. 1B). This occurs because the hazard ratio remains constant over time; thus, every patient’s projected trajectory differs only by a scalar factor determined by the model.

**Figure 1.**
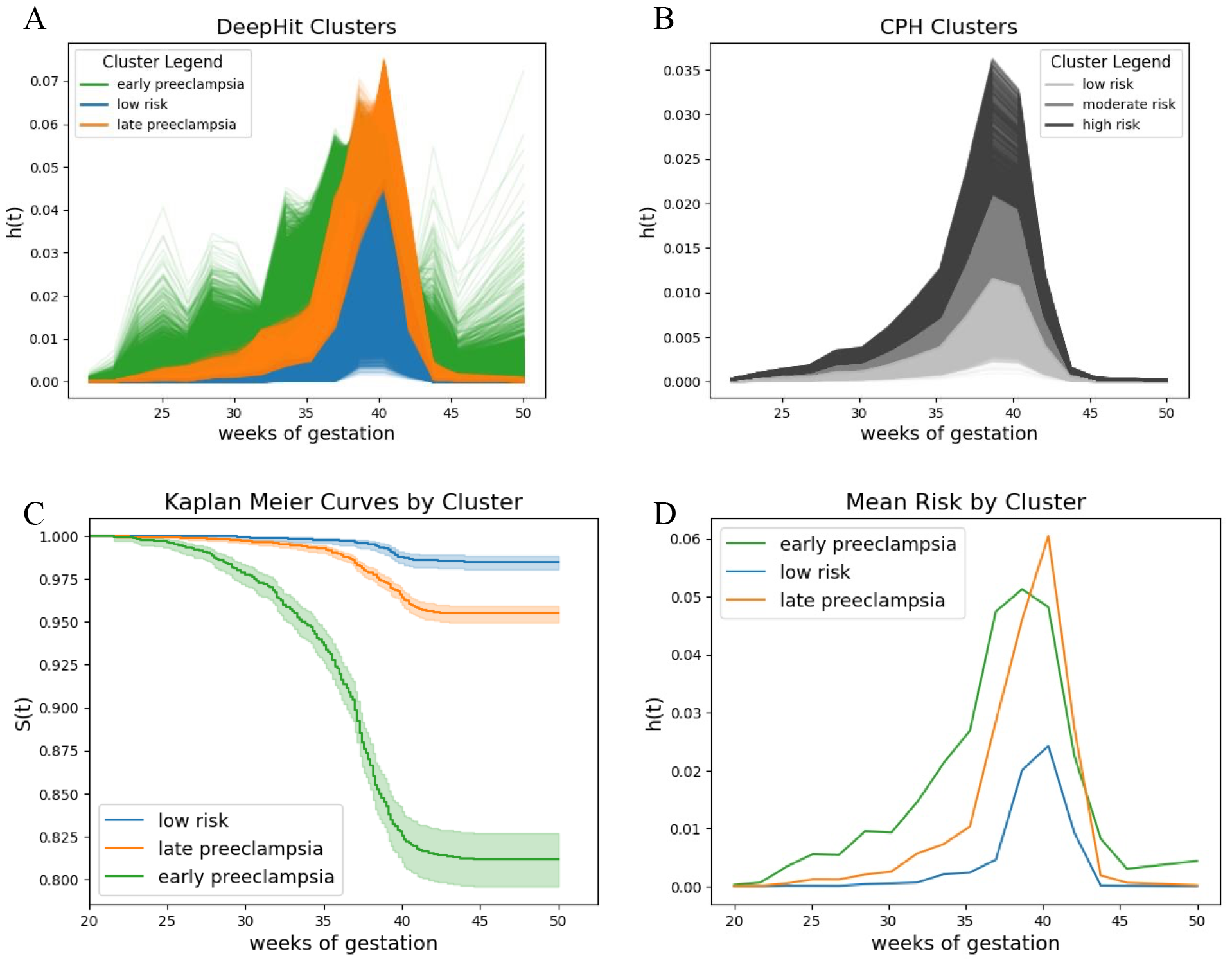
Results from the deep survival analysis. A) Time series k-means clustering with DeepHit B) Time series k-means clustering with Cox Proportional Hazards model C) Kaplan Meier survival by DeepHit clusters D) Average preeclampsia risk function by DeepHit clusters. Abbreviations, CPH: Cox Proportional Hazards model

### 3.3 Feature Importance

Leveraging the outcomes of our clustering analysis, we employed SHAP to determine the highest-ranking features that influence the allocation of patients to distinct clusters. Fig 2 shows that top features are predominantly associated with blood pressure, demographic, and medical history variables across all clusters.

**Figure 2.**
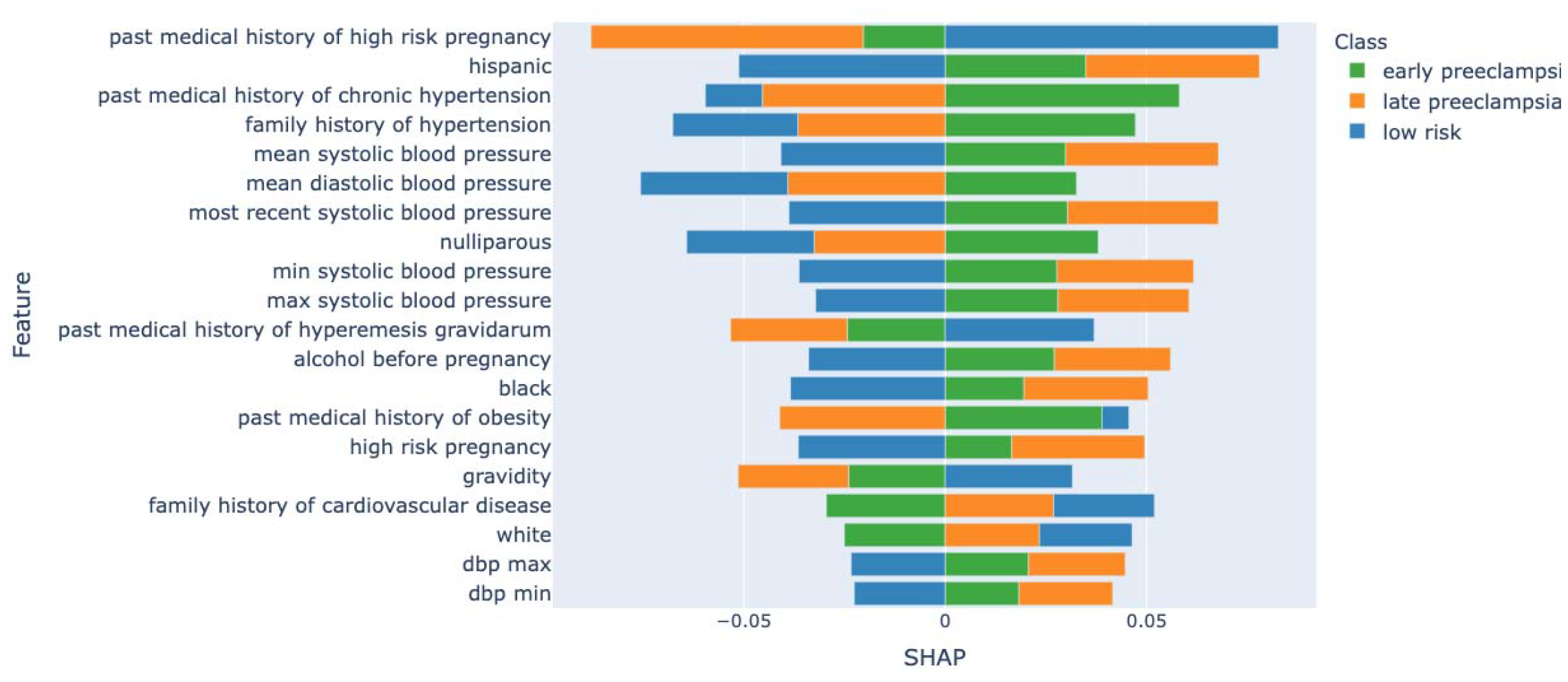
Mean SHAP values by feature for DeepHit clusters

Individual clusters mirror the overall trends, demonstrating consistency in the importance of blood pressure and relevant demographic and medical history across all clusters. Certain individual features attain heightened importance within specific clusters, exemplified by instances such as the pronounced impact of the medical history of chronic hypertension and race, as shown in Fig 3. A, B, and C.

**Figure 3.**
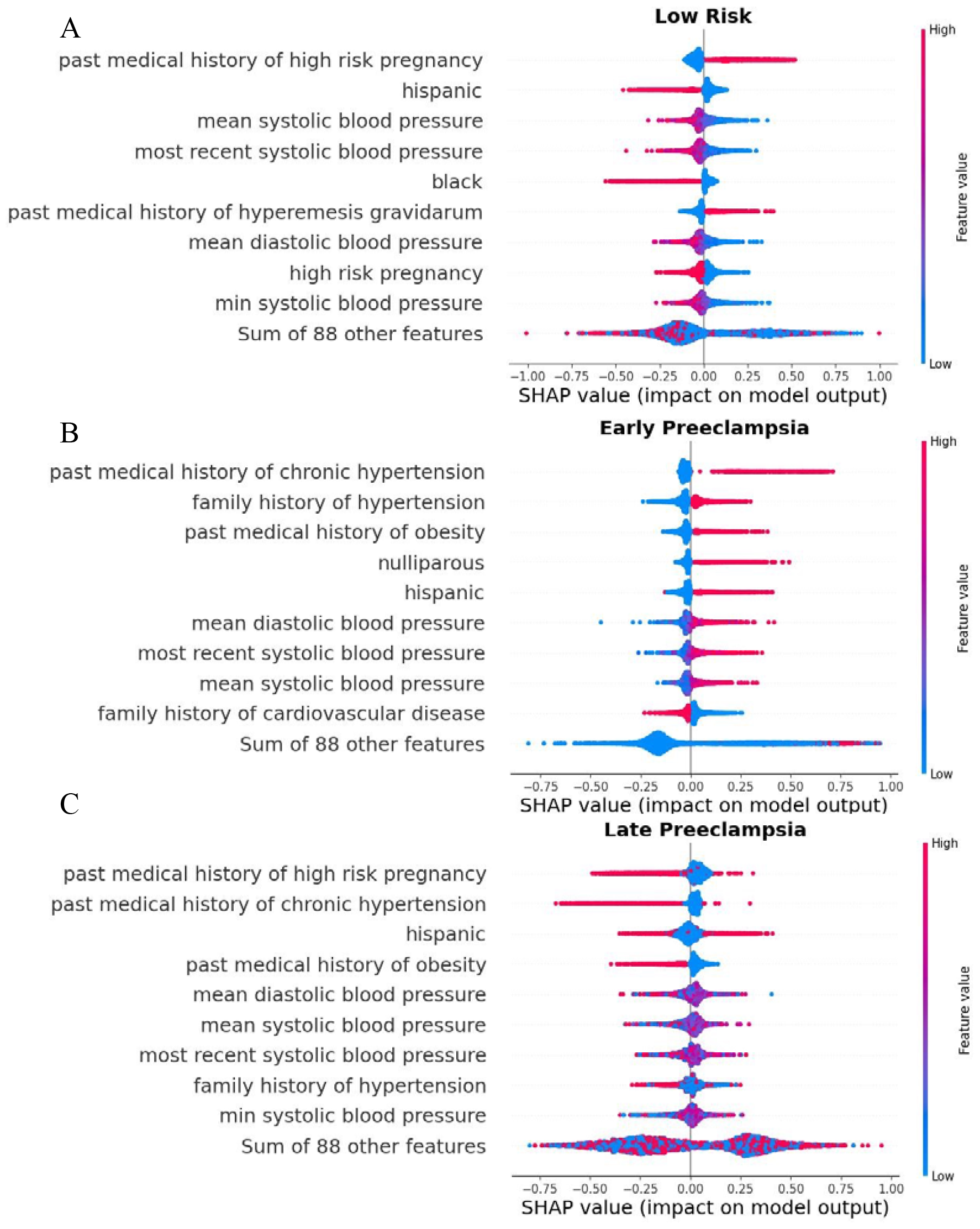
Interpretability analysis. A) SHAP values for low risk DeepHit cluster B) SHAP values for early preeclampsia DeepHit cluster C) SHAP values for late preeclampsia DeepHit cluster.

Figure 4 involves a systematic grouping of features based on the type of information they represent. This analysis sheds light on the relative contributions of each group of variables (medical history, demographics, vital signs, and laboratory results) within each cluster. Most notably, risk factors from medical history are more predictive of early-onset preeclampsia, while vital signs have increased relative significance in predicting late-onset preeclampsia and the low-risk groups.

**Fig. 4.**
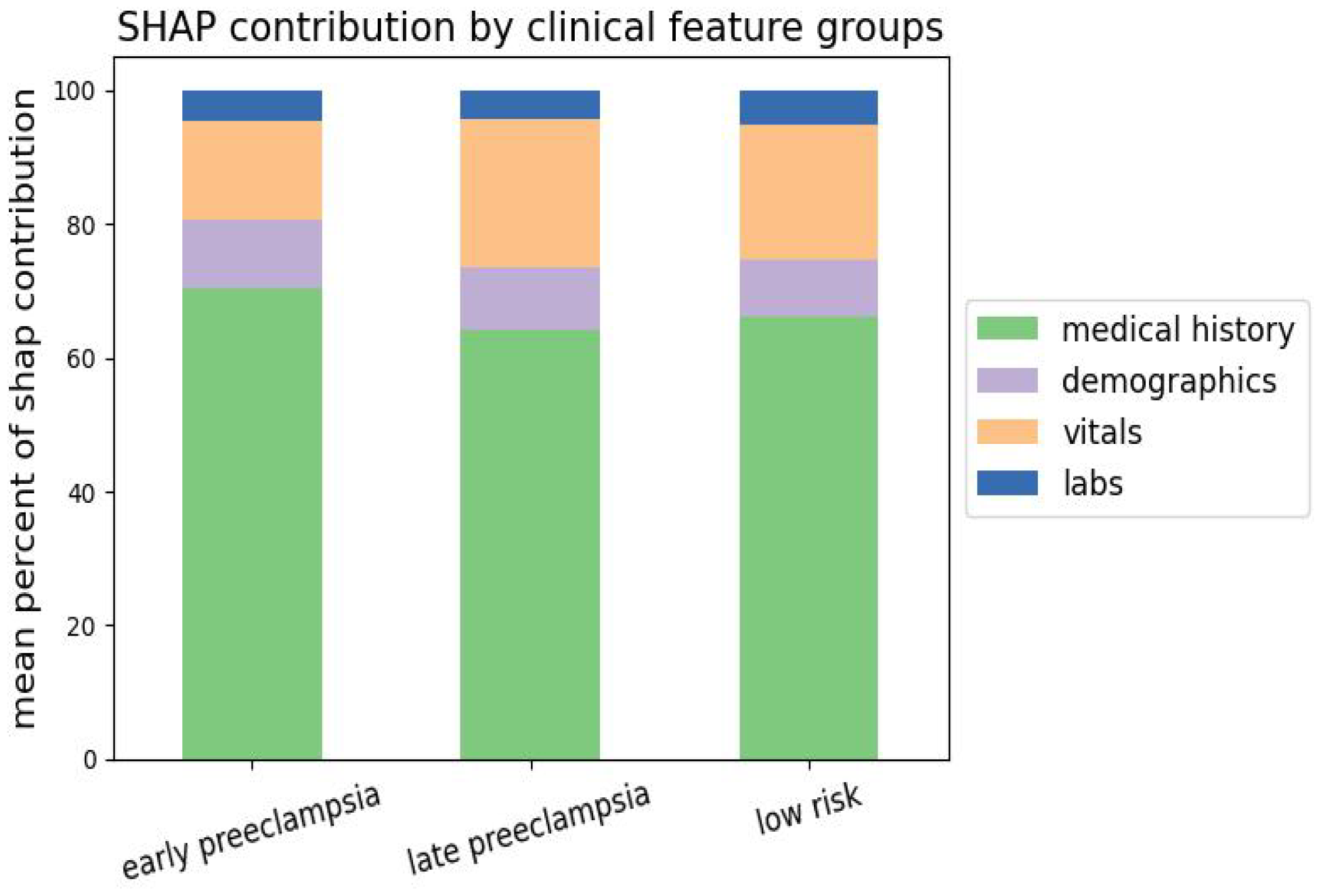
Average SHAP contribution of clinical feature groups

## 4. Discussion

In this study, we utilized nonparametric survival analysis, specifically the DeepHit model, to unravel the time-varying characteristics of a common pregnancy-associated condition, preeclampsia. DeepHit is particularly well-suited to this situation compared to traditional Cox regressions given the substantial variation in the risks associated with this condition over time. We found that DeepHit demonstrates comparable survival analysis metrics to the baseline Cox regression, achieving a concordance index (Ctd) of 0.84 and an AUCfinal of 0.78, facilitating meaningful comparisons with traditional classification methods. In contrast with traditional methods, DeepHit can capture evolving temporal shifts, underscoring its effectiveness in predicting personalized risk trajectories. Through temporal clustering, the study identifies three distinct risk patterns: low-risk, early-onset preeclampsia, and late-onset preeclampsia. The incorporation of SHAP values provides insights into the features, including blood pressures and medical comorbidities, that influence the formation of these clusters, offering a comprehensive understanding of the factors shaping patient risk evolution. The time-varying complication risk is a phenomenon found in many domains of medicine.

By framing the problem as a survival analysis, we capitalize on several advantages, specifically in navigating the intricate temporal complexities associated with preeclampsia. In contrast to models reliant on restrictive parametric assumptions, our methodology captures the relationships among gestational age, delivery, and disease risk. This is particularly crucial given the clinical variations observed across antepartum, intrapartum, and postpartum presentations. This research both emphasizes the limitations inherent in proportional hazards’ assumptions and accentuates the clinical relevance of employing nonparametric approaches to enhance the precision of risk assessment.

The DeepHit model’s ability to handle high dimensionality and navigate complex relationships among covariates is paramount in addressing preeclampsia risk. This becomes especially crucial given the multifaceted nature of the condition and the myriad clinical factors influencing risk across different stages. Our findings substantiate DeepHit’s competent performance in modeling intricate relationships between covariates and risk, particularly within complex temporal patterns, as exemplified in preeclampsia.

### 4.1 Advantages of this Approach

The nonparametric approach, exemplified by the DeepHit model, introduces several advantages in modeling preeclampsia risk dynamics.

#### 4.1.1 Individual Trajectories

One key advantage lies in the model’s ability to model separate risk trajectories for individual patients. Unlike traditional methods such as the Cox proportional hazards model, DeepHit does not assume proportional hazards, allowing it to capture unique risk evolutions. This feature is particularly advantageous in understanding the distinct patterns within patient groups over time, as demonstrated in the study’s temporal clustering analysis, which provides insights into evolving risk profiles. Other approaches have attempted to differentiate these groups using separate models with an artificial division of patients into early and late groups[15]. However, this approach necessitates the selection of a timepoint for data partitioning, thereby imposing an artificial decision boundary. In contrast, DeepHit exhibits flexibility in modeling diverse risk profiles through an intrinsic and data-driven approach. It optimizes its diagnostic capability and the accurate ordering of patients based on the cumulative incidence function and log-likelihood losses. Unlike previous approaches that explicitly designate patient categories, DeepHit allows the model to discover insights within the data, enhancing its ability to discern patterns without predefined constraints.

#### 4.1.2 Predictive Capability

DeepHit has a strong performance in predicting preeclampsia risk, as demonstrated by metrics such as Ctd, AUCtd, and Brier scores that are comparable to the baseline Cox regression. In addition to DeepHit’s primary strengths in properties such as capturing nuanced temporal shifts and individualized risk trajectories, it exhibits robust predictive capabilities for overall risk using AUCfinal, showcasing comparable performance to both Cox proportional hazards model and leading classification models at 20 weeks of gestation[32].

#### 4.1.3 Features Responsible for Risk Timing

DeepHit also facilitates a granular understanding of the features responsible for the timing of preeclampsia risk. Based on feature importance analysis by SHAP values, the study identifies critical determinants that define risk categories. The overarching trends underscore the pivotal impact of these features in forming the patient clusters, reaffirming the essential role of blood pressure dynamics[33] and pertinent patient background information[34] in determining disease timing. This detailed insight into feature contributions enables clinicians to pinpoint the factors steering patient clusters, enhancing interpretability, and guiding targeted interventions. The associations of early-onset preeclampsia with chronic hypertension and nulliparity and late-onset preeclampsia with obesity and chronic hypertension have been well documented in multiple studies[15]. The relationship between Hispanic ethnicity and late-onset preeclampsia is novel and thus warrants further investigation. This highlights the potential of our approach to discover novel factors associated with time-varying risk. The adaptability of the nonparametric approach proves instrumental in discerning subtle variations in risk trajectories, ultimately improving the clinical utility of risk assessments for preeclampsia.

### 4.2 Clinical Applications

Using a nonparametric framework, this study provides an individualized risk assessment, enabling clinicians to tailor interventions based on the predicted onset of disease. The model’s ability to delineate risk trajectories unconstrained by proportional hazard assumptions allows for a more accurate representation of each patient’s journey. The resulting individualized perspective enhances precision in risk predictions, empowering proactive and targeted management strategies, such as increased surveillance and aspirin prophylaxis, especially in patients at high risk for early-onset preeclampsia. By addressing the specific profile of prolonged risk, clinicians can tailor interventions to individual patient needs, ultimately improving both maternal and fetal outcomes in the challenging context of preeclampsia.

The study’s temporal clustering analysis further reveals distinct risk patterns within patient groups over time. Clinicians can leverage this information for risk stratification, developing care plans adapted to specific temporal dynamics. Understanding critical features influencing the timing of preeclampsia risk, as identified through feature importance analysis, empowers clinicians to develop targeted intervention strategies.

This study also offers insight for future research in obstetric conditions. Exploring specific features influencing risk timing and developing targeted interventions based on these features can advance prenatal care practices, marking a significant step forward in maternal-fetal care. Embracing nonparametric models ensures a more accurate representation of risk evolution, contributing to improved decision-making. The adaptability and precision of the nonparametric approach have the potential to facilitate enhanced risk communication with patients, fostering a better understanding and active engagement in healthcare decisions.

### 4.3 Limitations

A limitation of deep survival models is their complexity and black-box nature. We overcome this limitation by applying clustering and interpretability approaches. The reliance of deep survival models on discrete time intervals is another consideration. Nonparametric models often require the event of interest to be discretized into intervals. This discretization introduces a trade-off between temporal granularity and computational feasibility. Binning events into discrete time intervals may oversimplify the temporal nuances of preeclampsia progression, potentially leading to the loss of fine-grained information. Consequently, the model’s ability to precisely capture rapid changes or subtle shifts in risk within intervals may be constrained.

The generalizability of our model to diverse populations and healthcare contexts warrants careful consideration. While our study demonstrates robust performance within the specific dataset used for training and validation, external validation on independent datasets representing varied patient demographics and healthcare settings is essential to ensure the model’s reliability and applicability across different populations. In addition, deep survival models are computationally intensive, and hold promise with large, multi-dimensional datasets.

Despite these limitations, our study demonstrates the great potential of enhanced models and provides clinically meaningful insights. The innovative application of nonparametric models to unravel the temporal dynamics of preeclampsia risk represents a novel approach to preeclampsia risk prediction. The nuanced insights gained into the evolving nature of the disorder, coupled with the transparent interpretability provided by SHAP values, offer valuable contributions to both clinicians and researchers.

## 5. Conclusions

We evaluated deep survival analysis for predicting the time-varying risk of preeclampsia during pregnancy and postpartum. This technique demonstrates significant advantages over traditional models. Our results demonstrate the superiority of DeepHit over Cox models in capturing intricate temporal patterns and dynamic relationships between covariates, particularly in handling high-dimensional data and evolving risk hazards over time. The analysis reveals that the risk for preeclampsia is time-varied, leading to the identification of distinct risk trajectories, low-risk, early-onset preeclampsia, and late-onset preeclampsia groups, each associated with distinct risk factors. The time series k-means clustering further illustrates the improved ability of DeepHit to delineate diverse disease risk trajectories, providing valuable insights for early and individualized intervention. We highlight the potential of deep survival models in personalized risk trajectory prediction, offering a more accurate and flexible framework for understanding complex temporal dynamics in the context of maternal health.

## Data Availability

The patient data used in this study contains identifiable protected health information and therefore cannot be shared publicly. MGB investigators with an appropriate IRB approval can contact the corresponding author directly regarding data access.

## ABBREVIATIONS

AUC: area under the curve
BMI: body mass index
CIF: cumulative incidence function
CPH: Cox proportional hazards model
Ctd: time-dependent Concordance Index
DBP: diastolic blood pressure
IBS: Integrated Brier Score
SBP: systolic blood pressure
SHAP: SHapley Additive exPlanations

## Acknowledgments

KJG reports funding from NIH/NHLBI grants K08 HL146963, K08 HL146963-02S1, and R03 HL162756. VPK reports funding from the NIH/NHLBI grants 1K08HL161326-01A1, Anesthesia Patient Safety Foundation (APSF), and BWH IGNITE Award.

## References

[1] S.M. Ahmad, N.M. Ahmed, Classification based on event in survival machine learning analysis of cardiovascular disease cohort, BMC Cardiovasc. Disord. 23 (2023) 310. 10.1186/s12872-023-03328-2.

[2] J. Shen, L. Wang, S. Daignault, D.E. Spratt, T.M. Morgan, J.M.G. Taylor, Estimating the Optimal Personalized Treatment Strategy Based on Selected Variables to Prolong Survival via Random Survival Forest with Weighted Bootstrap, J. Biopharm. Stat. 28 (2018) 362–381. 10.1080/10543406.2017.1380036.

[3] J. Le-Rademacher, X. Wang, Time-To-Event Data: An Overview and Analysis Considerations, J. Thorac. Oncol. Off. Publ. Int. Assoc. Study Lung Cancer. 16 (2021) 1067–1074. 10.1016/j.jtho.2021.04.004.

[4] D.R. Cox, Regression Models and Life-Tables, J. R. Stat. Soc. Ser. B Methodol. 34 (1972) 187–202. 10.1111/j.2517-6161.1972.tb00899.x.

[5] H. Kvamme, Ø. Borgan, I. Scheel, Time-to-Event Prediction with Neural Networks and Cox Regression, (2019). 10.48550/arXiv.1907.00825.

[6] X. Zhu, J. Yao, J. Huang, Deep convolutional neural network for survival analysis with pathological images, in: 2016 IEEE Int. Conf. Bioinforma. Biomed. BIBM, 2016: pp. 544–547. 10.1109/BIBM.2016.7822579.

[7] CapSurv: Capsule Network for Survival Analysis With Whole Slide Pathological Images | IEEE Journals & Magazine | IEEE Xplore, (n.d.). https://ieeexplore.ieee.org/document/8651474 (accessed November 30, 2023).

[8] P. Kopper, S. Wiegrebe, B. Bischl, A. Bender, D. Rügamer, DeepPAMM: Deep Piecewise Exponential Additive Mixed Models for Complex Hazard Structures in Survival Analysis, (2022). 10.48550/arXiv.2202.07423.

[9] A. Bennis, S. Mouysset, M. Serrurier, DPWTE: A Deep Learning Approach to Survival Analysis Using a Parsimonious Mixture of Weibull Distributions, in: I. Farkaš, P. Masulli, S. Otte, S. Wermter (Eds.), Artif. Neural Netw. Mach. Learn. – ICANN 2021, Springer International Publishing, Cham, 2021: pp. 185–196. 10.1007/978-3-030-86340-1_15.

[10] C. Lee, W. Zame, J. Yoon, M. van der Schaar, DeepHit: A Deep Learning Approach to Survival Analysis With Competing Risks, Proc. AAAI Conf. Artif. Intell. 32 (2018). 10.1609/aaai.v32i1.11842.

[11] E.A.P. Steegers, P. von Dadelszen, J.J. Duvekot, R. Pijnenborg, Pre-eclampsia, Lancet Lond. Engl. 376 (2010) 631–644. 10.1016/S0140-6736(10)60279-6.

[12] Gestational Hypertension and Preeclampsia: ACOG Practice Bulletin, Number 222, Obstet. Gynecol. 135 (2020) e237–e260. 10.1097/AOG.0000000000003891.

[13] S. Khan, A.B. Siddique, S. Jabeen, A.T. Hossain, M.M. Haider, F.T. Zohora, M.M. Rahman, S. El Arifeen, A.E. Rahman, K. Jamil, Preeclampsia and eclampsia-specific maternal mortality in Bangladesh: Levels, trends, timing, and care-seeking practices, J. Glob. Health. 13 (n.d.) 07003. 10.7189/jogh.13.07003.

[14] A.B. Wallis, A.F. Saftlas, J. Hsia, H.K. Atrash, Secular trends in the rates of preeclampsia, eclampsia, and gestational hypertension, United States, 1987-2004, Am. J. Hypertens. 21 (2008) 521–526. 10.1038/ajh.2008.20.

[15] S. Lisonkova, K.S. Joseph, Incidence of preeclampsia: risk factors and outcomes associated with early- versus late-onset disease, Am. J. Obstet. Gynecol. 209 (2013) 544.e1-544.e12. 10.1016/j.ajog.2013.08.019.

[16] R.G. Sinkey, A.N. Battarbee, N.A. Bello, C.W. Ives, S. Oparil, A.T. Tita, Prevention, Diagnosis and Management of Hypertensive Disorders of Pregnancy: A Comparison of International Guidelines, Curr. Hypertens. Rep. 22 (2020) 66. 10.1007/s11906-020-01082-w.

[17] Low-Dose Aspirin Use During Pregnancy, (n.d.). https://www.acog.org/clinical/clinical-guidance/committee-opinion/articles/2018/07/low-dose-aspirin-use-during-pregnancy (accessed November 25, 2023).

[18] S.S. Aljameel, M. Alzahrani, R. Almusharraf, M. Altukhais, S. Alshaia, H. Sahlouli, N. Aslam, I.U. Khan, D.A. Alabbad, A. Alsumayt, Prediction of Preeclampsia Using Machine Learning and Deep Learning Models: A Review, Big Data Cogn. Comput. 7 (2023) 32. 10.3390/bdcc7010032.

[19] M. Liu, X. Yang, G. Chen, Y. Ding, M. Shi, L. Sun, Z. Huang, J. Liu, T. Liu, R. Yan, R. Li, Development of a prediction model on preeclampsia using machine learning-based method: a retrospective cohort study in China, Front. Physiol. 13 (2022). https://www.frontiersin.org/articles/10.3389/fphys.2022.896969 (accessed November 30, 2023).

[20] H. Sufriyana, Y.-W. Wu, E.C.-Y. Su, Artificial intelligence-assisted prediction of preeclampsia: Development and external validation of a nationwide health insurance dataset of the BPJS Kesehatan in Indonesia, EBioMedicine. 54 (2020) 102710. 10.1016/j.ebiom.2020.102710.

[21] D. Wright, M.Y. Tan, N. O’Gorman, L.C. Poon, A. Syngelaki, A. Wright, K.H. Nicolaides, Predictive performance of the competing risk model in screening for preeclampsia, Am. J. Obstet. Gynecol. 220 (2019) 199.e1-199.e13. 10.1016/j.ajog.2018.11.1087.

[22] B.M. Sibai, Etiology and management of postpartum hypertension-preeclampsia, Am. J. Obstet. Gynecol. 206 (2012) 470–475. 10.1016/j.ajog.2011.09.002.

[23] S. Tran, J. Fogel, S. Karrar, P. Hong, Comparison of process outcomes, clinical symptoms and laboratory values between patients with antepartum preeclampsia, antepartum with persistent postpartum preeclampsia, and new onset postpartum preeclampsia, J. Gynecol. Obstet. Hum. Reprod. 49 (2020) 101724. 10.1016/j.jogoh.2020.101724.

[24] B. George, S. Seals, I. Aban, Survival analysis and regression models, J. Nucl. Cardiol. Off. Publ. Am. Soc. Nucl. Cardiol. 21 (2014) 686–694. 10.1007/s12350-014-9908-2.

[25] R.Y. Cohen, V.P. Kovacheva, A Methodology for a Scalable, Collaborative, and Resource-Efficient Platform, MERLIN, to Facilitate Healthcare AI Research, IEEE J. Biomed. Health Inform. 27 (2023) 3014–3025. 10.1109/JBHI.2023.3259395.

[26] K.P. Vatcheva, M. Lee, J.B. McCormick, M.H. Rahbar, Multicollinearity in Regression Analyses Conducted in Epidemiologic Studies, Epidemiol. Sunnyvale Calif. 6 (2016) 227. 10.4172/2161-1165.1000227.

[27] F.E. Harrell, R.M. Califf, D.B. Pryor, K.L. Lee, R.A. Rosati, Evaluating the yield of medical tests, JAMA. 247 (1982) 2543–2546.

[28] Optuna: A hyperparameter optimization framework — Optuna 3.1.0 documentation, (n.d.). https://optuna.readthedocs.io/en/stable/ (accessed March 13, 2023).

[29] Time-dependent ROC curve analysis in medical research: current methods and applications | BMC Medical Research Methodology | Full Text, (n.d.). https://bmcmedresmethodol.biomedcentral.com/articles/10.1186/s12874-017-0332-6 (accessed November 27, 2023).

[30] R. Tavenard, J. Faouzi, G. Vandewiele, F. Divo, G. Androz, C. Holtz, M. Payne, R. Yurchak, M. RuÃŸwurm, K. Kolar, E. Woods Tslearn, A Machine Learning Toolkit for Time Series Data, J Mach Learn Res. 21 (n.d.) 1–6. http://jmlr.org/papers/v21/20-091.html.

[31] A. Gramegna, P. Giudici, SHAP and LIME: An Evaluation of Discriminative Power in Credit Risk, Front. Artif. Intell. 4 (2021) 752558. 10.3389/frai.2021.752558.

[32] S. Li, Z. Wang, L.A. Vieira, A.B. Zheutlin, B. Ru, E. Schadt, P. Wang, A.B. Copperman, J.L. Stone, S.J. Gross, Y.-H. Kao, Y.K. Lau, S.M. Dolan, E.E. Schadt, L. Li, Improving preeclampsia risk prediction by modeling pregnancy trajectories from routinely collected electronic medical record data, Npj Digit. Med. 5 (2022) 1–16. 10.1038/s41746-022-00612-x.

[33] R. Fox, J. Kitt, P. Leeson, C.Y.L. Aye, A.J. Lewandowski, Preeclampsia: Risk Factors, Diagnosis, Management, and the Cardiovascular Impact on the Offspring, J. Clin. Med. 8 (2019) 1625. 10.3390/jcm8101625.

[34] K. Duckitt, D. Harrington, Risk factors for pre-eclampsia at antenatal booking: systematic review of controlled studies, BMJ. 330 (2005) 565. 10.1136/bmj.38380.674340.E0.

