## Supplemental Figures and Tables for "Deep Survival Analysis for Interpretable Time-Varying Prediction of Preeclampsia Risk"

Supplementary Information

| **Supplementary Table 1. Clinical Features and Groups** | |
| --- | --- |
| **Groups** | **Features** |
| Labs | Calcium serum mean, calcium serum max, creatinine serum range, RBC count rage, RBC count mean, RBC count max, RBC count min, WBC count range, WBC count mean, WBC count max, WBC count min, uric acid mean, platelets min, uric acid max, uric acid min, hypertensive medication |
| Medical History | Family history of (cardiovascular disease, diabetes, heart disease, hyperlipidemia, hypertension, preeclampsia, stroke), past medical history of (asthma, chronic hypertension, diabetes, gestational hypertension, high risk pregnancy, hyperemesis gravidarum, migraine, obesity, preeclampsia, pregnancy-related fatigue, lupus, preterm birth, autoimmune disease), alcohol use, drug use, cigarette use, anemia, headaches, oligohydramnios, high-risk pregnancy, hyperemesis gravidarum, pregnancy-related fatigue, hypertension, IVF, multiple gestation, nulliparous, proteinuria, gravidity, parity, interpregnancy interval |
| Demographics | Private insurance, public insurance, White, Black, Hispanic, maternal age |
| Vital Signs | BMI range, BMI last week range, BMI last week max, BMI last week mean, BMI last week min, BMI max, BMI mean, BMI min, BMI most recent, heart rate range, heart rate max, heart rate mean, heart rate min, heart rate last week range, heart rate last week mean, heart rate last week max, heart rate last week min, heart rate most recent, weight last week range, weight last week mean, weight last week max, weight last week min, weight range, weight mean, weight max, weight min, weight most recent, SBP range, SBP last week max, SBP last week min, SBP last week mean, SBP max, SBP max, SBP mean, SBP min, SBP most recent, DBP range, DBP last week max, DBP last week min, DBP last week mean, DBP max, DBP max, DBP mean, DBP min, DBP most recent |

Abbreviations. RBC, red blood cell, WBC, white blood cell, IVF, in vitro fertilization, BMI, body mass index, SBP, systolic blood pressure, DBP, diastolic blood pressure

**Supplementary Table 2. Data used for model training and testing**

| **Feature** | **Preeclampsia** | **No Preeclampsia** | **P value** |
| --- | --- | --- | --- |
| Self-Reported Race, White | 2380 (57 %) | 39980 (64 %) | < 0.01 |
| Self-Reported Race, Black | 815 (19 %) | 6651 (11 %) | < 0.01 |
| Self-Reported Ethnicity,  Hispanic | 855 (20 %) | 10504 (17 %) | < 0.01 |
| Maternal age, y | 33.4 (29.94 - 37.02) | 33.18 (30.21 - 36.13) | < 0.01 |
| Family history of heart disease | 91 (2 %) | 1264 (2 %) | 0.55 |
| Family history of hypertension | 3205 (76 %) | 42654 (69 %) | < 0.01 |
| Family history of preeclampsia | 105 (3 %) | 1732 (3 %) | 0.29 |
| Any history of alcohol use | 1304 (31 %) | 19075 (31 %) | 0.65 |
| High risk pregnancy | 2508 (60 %) | 30997 (50 %) | < 0.01 |
| Hyperemesis gravidarum | 325 (8 %) | 3907 (6 %) | < 0.01 |
| Pregnancy related fatigue | 525 (13 %) | 7031 (11 %) | 0.025 |
| Drugs of abuse | 282 (7 %) | 2634 (4 %) | < 0.01 |
| Chronic hypertension | 798 (19 %) | 2030 (3 %) | < 0.01 |
| Antihypertensive medication | 311 (7 %) | 408 (1 %) | < 0.01 |
| Nulliparous | 827 (20 %) | 9378 (15 %) | < 0.01 |
| Chronic proteinuria | 120 (3 %) | 64 (0 %) | < 0.01 |
| Gravidity | 2.0 (1.0 - 3.0) | 2.0 (1.0 - 3.0) | 0.73 |
| Parity | 1.0 (0.0 - 2.0) | 1.0 (1.0 - 2.0) | 0.045 |
| Interpregnancy interval, y | 2.41 (1.81 - 3.2) | 2.3 (1.81 - 3.02) | < 0.01 |
| Past history of preterm birth | 0.0 (0.0 - 1.0) | 0.0 (0.0 - 0.0) | < 0.01 |
| Labs rbc count diff | 0.99 (0.68 - 1.35) | 0.7 (0.47 - 1.0) | < 0.01 |
| Labs rbc count mean | 3.92 (3.66 - 4.17) | 3.96 (3.75 - 4.18) | < 0.01 |
| Labs rbc count max | 4.4 (4.16 - 4.65) | 4.31 (4.1 - 4.55) | < 0.01 |
| Labs rbc count min | 3.43 (3.03 - 3.74) | 3.61 (3.31 - 3.87) | < 0.01 |
| Anemia | 673 (16 %) | 9095 (15 %) | 0.021 |
| Autoimmune history | 503 (12 %) | 3558 (6 %) | < 0.01 |
| History of smoking | 628 (15 %) | 7023 (11 %) | < 0.01 |
| Chronic headaches | 830 (20 %) | 9218 (15 %) | < 0.01 |
| Past medical history of diabetes | 512 (12 %) | 2824 (5 %) | < 0.01 |
| Past medical history of systemic lupus erythematosus | 57 (1 %) | 478 (1 %) | < 0.01 |
| SBP diff, mmHg | 78 (65 - 90) | 49 (40 - 59) | < 0.01 |
| SBP max, mmHg | 175 (163 - 185) | 140 (131 - 149) | < 0.01 |
| SBP mean, mmHg | 132 (127 - 138) | 113 (108 - 119) | < 0.01 |
| SBP min, mmHg | 98 (90 - 102) | 90 (86 - 97) | < 0.01 |
| SBP most recent, mmHg | 124(116 - 132) | 112 (104 - 121) | < 0.01 |
| DBP diff, mmHg | 52 (44 - 61) | 37 (31 - 44) | < 0.01 |
| DBP max, mmHg | 104 (98 - 111) | 86 (81 - 93) | < 0.01 |
| DBP mean, mmHg | 77 (73 - 81) | 66 (62 - 70) | < 0.01 |
| DBP min, mmHg | 53 (48 - 56) | 50 (46 - 53) | < 0.01 |
| DBP most recent, mmHg | 79 (70 - 84) | 70 (62 - 76) | < 0.01 |
| BMI max, kg/m^2^ | 33.5 (29.4 - 38.9) | 29.8 (26.8 - 33.7) | < 0.01 |
| BMI mean, kg/m^2^ | 31.3 (27.2 - 36.58) | 27.71 (24.86 - 31.6) | < 0.01 |
| BMI min, kg/m^2^ | 28.2 (24.0 - 33.8) | 24.8 (22.0 - 29.0) | < 0.01 |
| BMI most recent, kg/m^2^ | 33.2 (29.1 - 38.44) | 29.5 (26.6 - 33.4) | < 0.01 |
| Heart rate diff, bpm | 51 (39 - 65) | 44 (32 - 59) | < 0.01 |
| Heart rate max, bpm | 112 (101 - 125) | 107 (96 - 120) | < 0.01 |
| Heart rate mean, bpm | 83 (77 - 90) | 81 (75 - 88) | < 0.01 |
| Heart rate min, bpm | 61 (55 - 68) | 62 (56 - 68) | < 0.01 |
| Heart rate most recent, bpm | 80 (74 - 89) | 77.0 (70.0 - 84.0) | < 0.01 |
| Weight max, kg | 88.13 (77.11 - 103.42) | 79.56 (71.21 - 90.26) | < 0.01 |
| Weight mean, kg | 82.06 (71.25 - 97.49) | 73.94 (66.04 - 84.49) | < 0.01 |
| Weight min, kg | 73.75 (63.05 - 89.8) | 65.95 (58.42 - 76.66) | < 0.01 |
| Weight most recent, kg | 87.54 (76.66 - 102.51) | 79.11 (70.76 - 89.81) | < 0.01 |
| Outcome: Preeclampsia bool | 4196 (100 %) | 0 (0 %) | < 0.01 |
| Outcome: No preeclampsia bool | 0 (0 %) | 62194 (100 %) | < 0.01 |
| Abbreviations. RBC, red blood cell, BMI, body mass index, SBP, systolic blood pressure, DBP, diastolic blood pressure | | | |

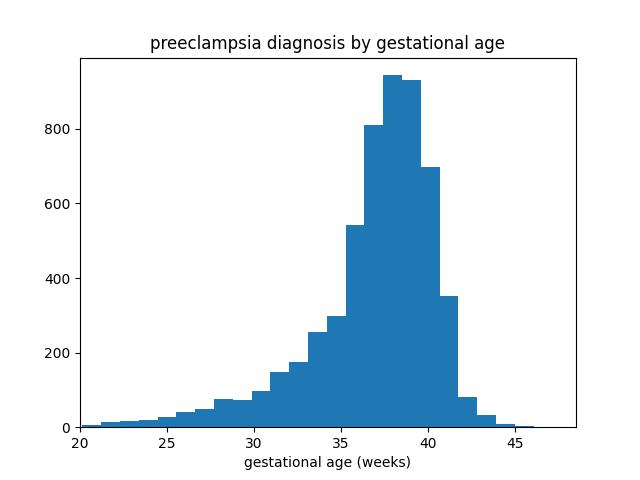

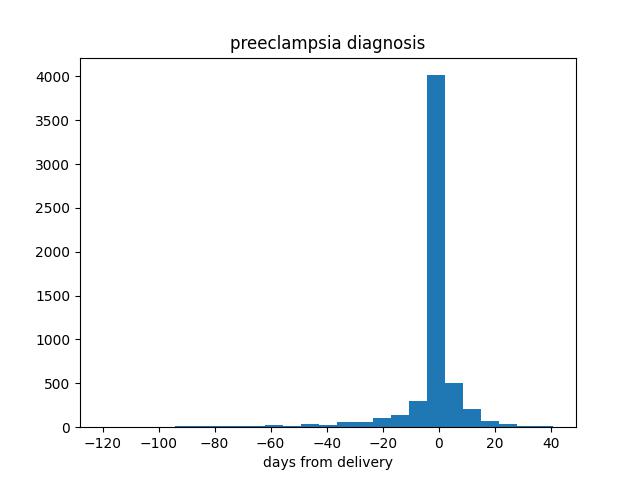
Supplementary Figure 1. Time to preeclampsia diagnosis. A. Histogram based on preeclampsia diagnosis relative to the delivery date. B. Histogram based on preeclampsia diagnosis relative to the gestational age.
